# Spatial Analysis Reveals Impaired Immune Cell Function within the Tumor Microenvironment of HIV-associated Non-small Cell Lung Cancer

**DOI:** 10.1101/2023.10.21.23297229

**Authors:** Shruti Desai, Syim Salahuddin, Ramsey Yusuf, Kishu Ranjan, Jianlei Gu, Ya-Wei Lin, Ronen Talmon, Yuval Kluger, Hongyu Zhao, Kurt Schalper, Brinda Emu

## Abstract

**Background:** Among people with HIV (PWH), lung cancer is the leader cause of cancer mortality, with increased risk and poor clinical outcomes compared to people without HIV (PWOH). HIV is known to result in persistent global immune dysfunction despite antiretroviral therapy, but little is known about the lung cancer tumor microenvironment. This study explored whether the tumor microenvironment (TME) of HIV-associated non-small cell lung cancer (NSCLC) is associated with an immunoregulator environment that limits tumor-specific immune responses.

**Methods:** A tissue microarray was constructed with NSCLC tumors from 18 PWH and 19 PWOH (matched for histological subtype, stage, year of diagnosis, age, sex and smoking status), and incubated with metal-conjugated antibodies for evaluation by imaging mass cytometry (IMC). IMC marker scores were extracted by automated cell segmentation and single-cell data was analyzed by Phenograph using unsupervised cell-segmentation and clustering of cells. Evaluation of tumor infiltrating immune cells, CD4+ and CD8+ T cells as well as CD68+ tumor associated macrophages were characterized for marker expression using a linear mixed-effects model. Additionally, a computational strategy based on the PageRank mathematical algorithm was used in order to establish an unsupervised and cell segmentation-independent signature associated with HIV status to discriminate differential expression of immune cell markers within the TME of the two groups. Peripheral blood mononuclear cells (PBMCs) from HLA-A02 donors (PWH and PWOH) were co-incubated with HLA-A02 lung cancer cell lines to quantify tumor killing (by Annexin V staining) and expression of T cell markers Lag-3 and CD25.

**Results:** Within the TME from HIV+ tumors, there is comparable level of infiltration of lymphocytes and tumor associated macrophages (TAMs) compared to non-HIV tumors, with a trend towards increased CD8+ T cells and decreased CD4:CD8 ratio among HIV+ tumors. Using a random effects model of individual markers, HIV+ tumors revealed increased expression of Ki67 and Granzyme B (GRZB) among CD8+ T cells; increased Ki67 and PD-1 among CD4+ T cells; and increased PD-L1, PD-L2, and Ki67 among TAMS. Unsupervised clustering analysis from IMC data demonstrated differential distribution of tumor infiltrating CD8+ T cell clusters between HIV+ and non-HIV tumors, defined by marker expression patterns. Three clusters were significantly elevated in HIV+ tumors (57.1% vs. 21.7% in non-HIV tumors, p<0.0001). All three clusters had comparatively elevated PD-1 and Lag-3 expression with varying expression of activation and proliferation markers CD25 and Ki67. Within tumor-infiltrating CD4+ T cells, a cluster characterized by checkpoint protein expression (PD-1+ and LAG-3) was also highly represented in HIV+ cases (35.2% vs. 9.8% in non-HIV cases, p<0.0001). HIV+ tumor-associated macrophages (TAM) had higher expression of immunoregulatory molecules (PD-L1, PD-L2, B7-H3, B7-H4, IDO1 and VISTA), confirmed by the expansion of three clusters comprising 58.8% of TAMs vs. 17.8% in non-HIV tumors (p<0.0001). Discrimination of cells between HIV+ and HIV-TME was further confirmed by spectral graph theory with 84.6% accuracy, with a combination of markers on TAMs and T cells. Lastly, PBMCs from PWH exhibited decreased tumor killing when exposed to HLA-matched NSCLC cell lines compared to PBMCs from PWOH. CD8+ T cells from PWH additionally had increased expression of immune checkpoint inhibitor Lag-3 upon exposure to tumor cells.

**Conclusions:** Our study demonstrates that the TME of HIV+ patients is characterized by a unique immune landscape, distinct from that of PWOH, with evidence of expansion of immune cells with enhanced immunoregulatory phenotypes and associated with impaired anti-tumor responses.

## Introduction

Non-AIDS defining cancers (NADCs) have overtaken AIDS-defining malignancies as the leading cause of cancer-related deaths among people with HIV (PWH) in the United States. Lung cancer has emerged as the most common NADC, and a leading cause of overall mortality among PWH [1]. As many as 40% of all cancer deaths occur as a result of lung cancer, as well as 5% of all deaths in PWH [2, 3]. The overall risk of lung cancer has consistently been demonstrated to be greater in PWH compared with patients without HIV infection (PWOH), with as high as a three- to four-fold increase in risk, an effect independent of active combined antiretroviral treatment (cART), serum CD4+ T-cell count or CD4+ T-cell nadir [4–6]. Similar to PWOH, the vast majority of lung malignancies in PWH comprise the non-small cell lung cancer (NSCLC) subsets adenocarcinoma and squamous cell carcinoma, but they occur at a younger age and present with more advanced clinical stage than in the general population [1, 4, 7].

HIV infection results in depletion of CD4+ T-cells, predisposing PWH to opportunistic infections and AIDS-defining cancers. However, CD4+ T-cell depletion is not the sole immune perturbation seen in chronic HIV infection. Immune activation and global immune dysfunction are also hallmarks of HIV infection, phenomena only partially reversible with initiation and maintenance of cART [8]. It is currently unknown how global immune dysregulation in the setting of HIV infection affects local anti-tumor immune responses and/or tumor cell properties, and how this may impact prognosis and treatment-specific outcomes in patients with lung cancer.

The tumor immune microenvironment in NSCLC is a spatially complex array of multiple cell types including malignant epithelial cancer cells, non-malignant stromal cells, and diverse immune cell subsets with implications for prognosis and response to treatment [9–12]. The prominent role of local T-cell responses in tumor elimination is highlighted by the clinical success of therapies blocking key immune inhibitory receptors upregulated in antigen-activated T-cells such as CTLA-4 and the PD-1/PD-L1 pathways (collectively referred to as immune checkpoint blockers [ICBs]). However, most patients with lung cancer are immunocompetent and tumors are expected to develop in the presence of adaptive immune pressure. In PWH however, the normal interactions between a neoantigen and evolution of an effective immune response may be impaired.

Multiple studies have shown that the tumor microenvironment (TME) composition is a key determinant of cancer survival and sensitivity to ICBs. For instance, the level of local PD-L1 expression, density of tumor infiltrating lymphocytes (TILs), and functional profile of TILs are associated with clinical benefit to PD-1 axis blockers in patients with advanced NSCLC [9–12]. Using single-cell and spatially resolved analysis of human NSCLCs, we have previously characterized the local effector T-cell dysfunction and found that evidence of enhanced T-cell proliferation, terminal differentiation and upregulation of immune inhibitory receptors were associated with reduced sensitivity to PD-1 axis blockers among NSCLC patients [10–12]. Similar findings have been reported by others using single-cell RNA sequencing strategies [13]. Recent studies have also revealed a prominent role of TIL spatial location in the tumor bed and emphasized the relevance of spatial immune heterogeneity in sensitivity/resistance to PD-1 axis blockers in NSCLC [14]. In addition, an asymmetric distribution of TILs with the presence of cytotoxic T-cells in the periphery of tumors (e.g. “leading edge”) with a relative absence in the tumor center characterizes poorly immunogenic malignancies, which has been associated with unfavorable outcomes [15–17]. Collectively, these studies support a prominent role of TILs and the TME in NSCLC rejection and response to immunotherapy.

The expected impact of HIV on T cells within the TME of NSCLC remains unknown. Several aspects of the local immune dysfunction in NSCLC resemble those seen systemically in PWH, suggesting potentially additive or even synergistic defective consequences. In this first comprehensive study of the immunologic components of the TME among PWH and NSCLC, we conducted a comparative analysis of HIV-associated and clinically matched non-HIV associated NSCLC utilizing spatially resolved and multiplexed tissue imaging modalities. Here, we show that the TME of HIV-associated NSCLC has several features of immune exhaustion, increased T-cell proliferation, and increased tumor mutational burden, suggesting greater dysfunction in the context of HIV-associated NSCLC compared to HIV infection or NSCLC alone. We additionally show that circulating T cells from patients with HIV, compared to T cells from HLA-matched PWOH, demonstrate poor tumor killing and enhanced upregulation of T cell inhibitory receptors when exposed to lung tumor cell lines.

## Methods

### Patients, Samples and Tissue Microarray Construction

Formalin-fixed, paraffin-embedded (FFPE) NSCLC tissue from 18 patients with HIV infection who underwent biopsy/surgery at Yale New Haven Hospital between January 2001 and January 2016 were obtained from the archives of the Pathology Department at Yale University (New Haven, CT). NSCLC biopsies of an additional 19 patients without HIV infection served as controls and were matched based on major clinicopathologic factors including stage, histologic subtype, year of biopsy, age, smoking status, and gender. A tissue microarray (TMA) was constructed from 0.6 mm tissue cores of each FFPE sample with 2-fold redundancy as previously reported by our group [9–12, 14]. Lymph node, tonsil, and placental tissue were included in the TMA to serve as positive controls for TIL markers and PD-L1 signal. The collection and analysis of samples used in this study were approved by the Yale University Human Investigation Committee (HIC) protocol # 1608018220.

### Imaging Mass Cytometry (IMC)

Preparation of the TMA and antibody cocktail for IMC analysis was performed using a modified IMC Staining Protocol for FFPE Sections from Fluidigm. The TMA slide was first incubated at 60°C for 30 minutes in a slide oven; then incubated at room temperature for 20 minutes in fresh xylene under a fume hood for dewaxing twice; then hydrated with ethanol solutions of descending concentrations for 1 minute each. The TMA was then washed with tap water for 5 minutes on a shaker plate and incubated at 96°C in the antigen retrieval solution (1mM EDTA at a pH of 9) for 20 minutes, then cooled to 70°C for 10 minutes. The TMA was incubated in a blocking solution containing 0.3% bovine serum albumin (BSA) in TBS-T (Tween-20 0.05%) for 30 minutes at room temperature.

An antibody cocktail containing metal-conjugated antibodies (DNA-191/193Ir, Vimentin-143Nd, Tbet-145Nd, CD47-146Nd, pan-cytokeratin-148Nd, CD45Ro-149Sm, PDL1-150Nd, GAPDH 151Eu, B7H3-152Sm, LAG3-153Eu, TIM3-154Sm, FOXP3-155Gd, CD4-156Gd, B7-H4-158Gd, CD68-159Tb, PD1-160Gd, CD20-161Dy, CD8-162Dy, CD25-163Dy, VISTA-165Ho, Ki67-168Er, B2M-169Tm, CD3-170Er, IDO1-171Yb, PDL2-172Yb, Granzyme B-173Yb, Histone3-176Yb) diluted in 0.3% BSA in TBS-T was prepared. The TMA slide was incubated in antibody cocktail diluted in BSA 0.3%-TBS-T solution overnight at 4° C; washed twice with 0.05% TBS-T at room temperature on a shaker plate for 5 minutes each time; incubated for 30 minutes in a solution containing Intercalator-Ir191/Ir193 (dilution 1:2000 in TBS-T) for 60 minutes at room temperature; washed for 5 minutes in TBS-T twice, followed by a wash for 5 minutes in MQ grade ddH2O twice; and finally left to air-dry at room temperature for 30 minutes protected from humidity and sunlight. Data for IMC was acquired via Hyperion instrument (DVS: Fluidigm Sciences).

For IMC analysis, extraction of raw IMC marker score and development of tissue images was done using MCD Viewer v1.0.560.0 (Fluidigm). Unsupervised cell segmentation and mask generation was completed using Cell Profiler v3.1.8. Briefly, cell nuclei were identified as primary objects within Cell Profiler using positive DNA expression on Tagged Image File Format (TIFF) images created in MCD Viewer for each tissue sample. Cell segmentation was automated and based on distance approximation using a standard radius of 3 pixels from all previously identified primary objects (identified as nuclei by DNA intercalators and Histone3 staining). If two cells overlapped in area, cell boundaries were approximated using the median distance between their nuclei. The optimal distance threshold of 3 pixels was set after comparing cell segmentation results using a range of radii with manually identified cell boundaries on TIFF images. A mask image is generated, which outlines the boundaries of each unique cell identified via segmentation. Segmented cell masks from CellProfiler were extracted and imported into histoCAT (Histology Topography Cytometry Analysis Toolbox) v1.75 along with corresponding TIFF images for each marker of interest for each tissue sample. PhenoGraph was used for unsupervised clustering through the MATLAB-based tool CYT, and single cell data was extracted for analysis[18]. FlowJo v10.0.7 was used for gating of cell populations, identifying CK-CD68-CD20-CD3+CD4+CD8−, CK-CD68-CD20-CD3+CD4-CD8+, CK-CD68+CD20-CD3−, and CK+CD68-CD20-CD3-cell populations from IMC single cell data.

### Spectral Graph Theory

For every tumor case within the TMA, 14 patches of size 35μm² are selected and centered at the highest CK expression. For each patch, the “stack” of expressions across all antibodies is examined and using graph analysis, a feature vector is constructed. Then, the feature vectors of all patches from all cases are assembled into a representation matrix, where the rows correspond to the number of markers and the columns to the number of patches. This matrix is then subjected to a nonlinear dimensionality reduction using diffusion maps. Subsequently, the matrix in the reduced dimension is utilized to predict the HIV status. For the prediction, we use an RBF support vector machine (SVM) classifier and a leave-one-case-out cross-validation analysis. In this process, the classifier is trained with all the patches, except those from one case, and then, tested on the excluded patches.

### in vitro Tumor Killing Assay

A549 and PC9 cells were cultured in DMEM and RPMI1640 respectively, supplemented with 10% fetal bovine serum and antibiotics (10,000 U/mL penicillin, 10 µg/mL streptomycin), and incubated at 37°C and 5% CO_2_. Exponentially grown cells were used in this study. Cell lines were authenticated every 3–6 months according to laboratory protocols. A549 and PC9 cells were left unstimulated or stimulated with IFNγ and TNFα (20 ng/mL each) for 24 hours before being washed and co-cultured in the presence of PBMCS (at 2:1 or 5:1 ratio PBMC:tumor ratio) for 72 hours. PBMCs were obtained from HLA-02+ donors (PWH, n=3 and PWOH n=3). Post incubation, co-cultured cells were stained to analyze T cell markers, CD8 (BD, Cat# 566852; BD, Cat# 555368), CD25 (BD, Cat# 560987), CD69 (BD, Cat# 555530), LAG3 (Biolegend, Cat# 369306) and PD-1 (BD, Cat# 560795), and tumor cell markers EPCAM (Biolegend, Cat# 369306) and Annexin V (Biolegend, Cat# 640906) as per manufacturer instruction and analyzed by LSRII flow cytometer (BD Biosciences). Subsequent analysis was performed using FCS Express software version 7 (Devovo software).

### Statistical Analysis

The IMC signal from non-HIV and HIV+ cases were compared using Dunn’s multiple comparison test if statistical significance was calculated in a nonparametric Kruskal-Wallis test, unpaired t-tests, Mann Whitney test, or with Welch-correction and chi-square for continuous and categorical variables, respectively. For the Figures 3B,3E and 4B, statistical analyses were performed using a linear mixed-effects model, with individual effects treated as random effects. The significance threshold was set at 0.05, and the analyses were done using the lmerTest package (Version 3.1-3) in R Version 4.2.2. The difference in expression of annexin on tumor cells or Lag-3 on T cells in the functional assays was assessed using unpaired t tests, with adjustment for multiple comparisons.

## Results

### PWH and PWOH were matched for clinicopathologic characteristics

NSCLC tumor samples from 18 PWH and 19 PWOH were clinicopathologically matched and included in the analysis. Cancer histologic subtype, stage at cancer diagnosis, year of cancer diagnosis, age, gender and smoking status were comparable between the two groups (p>0.05; **Supplementary Table S1**). The median age among PWH and PWOH was 54 and 57, respectively. For PWH, the median CD4 count was 440, 77% were on cART at the time of cancer diagnosis, and 44% had an undetectable viral load (<400 copies/μL). The median time since HIV diagnosis was 17 years, and 44% had an AIDS diagnosis.

### Lymphocytes and Macrophages are able to infiltrate the Tumor Microenvironment of HIV-associated NSCLC

We studied the NSCLC samples using a standardized IMC panel for simultaneous visualization and spatially resolved measurement of 37 protein markers. These markers included cell-phenotype indicators (e.g. cytokeratin, CD3, CD4, CD8, CD68, CD-19), functional markers and potential candidate immunotherapy targets (e.g. GZB, PD-1, LAG-3, TIM-3, Ki-67, PD-L1, PD-L2, VISTA, IDO-1), and structural indicators for tissue compartment (Vimentin) and cell segmentation (Histone-3 and DNA intercalators). Representative images from the IMC panel staining are shown in **Figure 1A**. As expected, CD3+ TILs and CD68+ tumor-associated macrophages (TAMs) were predominantly located in the cytokeratin-negative stromal tissue areas and showed variable marker profiles. Tumor cells showed prominent cytokeratin positivity and focal expression of immunomodulatory markers such as β2-microglobulin, PD-L1 and PD-L2. After staining and acquisition, the tumor images from all cases were digitally segmented into single-cell populations using nuclear segmentation models and expression of phenotypic cell markers (**Figure 1B**).

**Figure 1.**
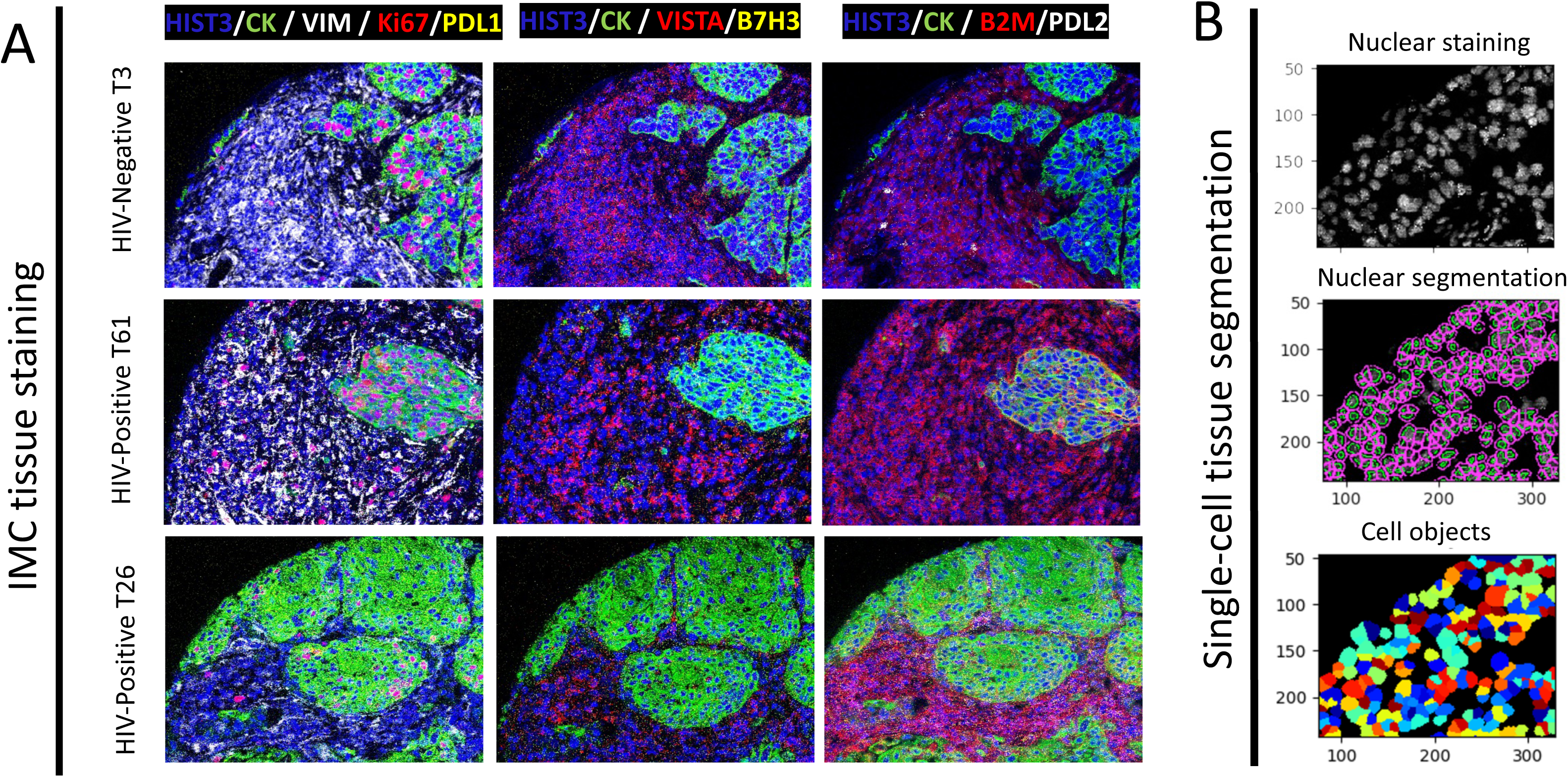
A) Representative multicolor microphotographs of a single NSCLC tumor stained with a 37-marker IMC panel. Captions of the same tumor displaying different markers as indicated in the color-code of each panel. Bar=100 um. B) Representative image showing the single cell segmentation of IMC tumor images using nuclear-cell markers and digital cell objects.

Quantification of main cell subpopulations of >35,000 individual cells obtained from cases/controls in the cohort showed comparable levels of CK+ tumor cells and CD68+ TAMs (**Figure 2A-B**) between PWH and PWOH. There is a trend towards lower CD4+ TILs (p=0.25), increased CD8+ TILs (p=0.08), and decreased CD4/CD8 T-cell ratio (p=0.14) among HIV+ NSCLCs, but the difference did not reach statistical significance.

**Figure 2.**
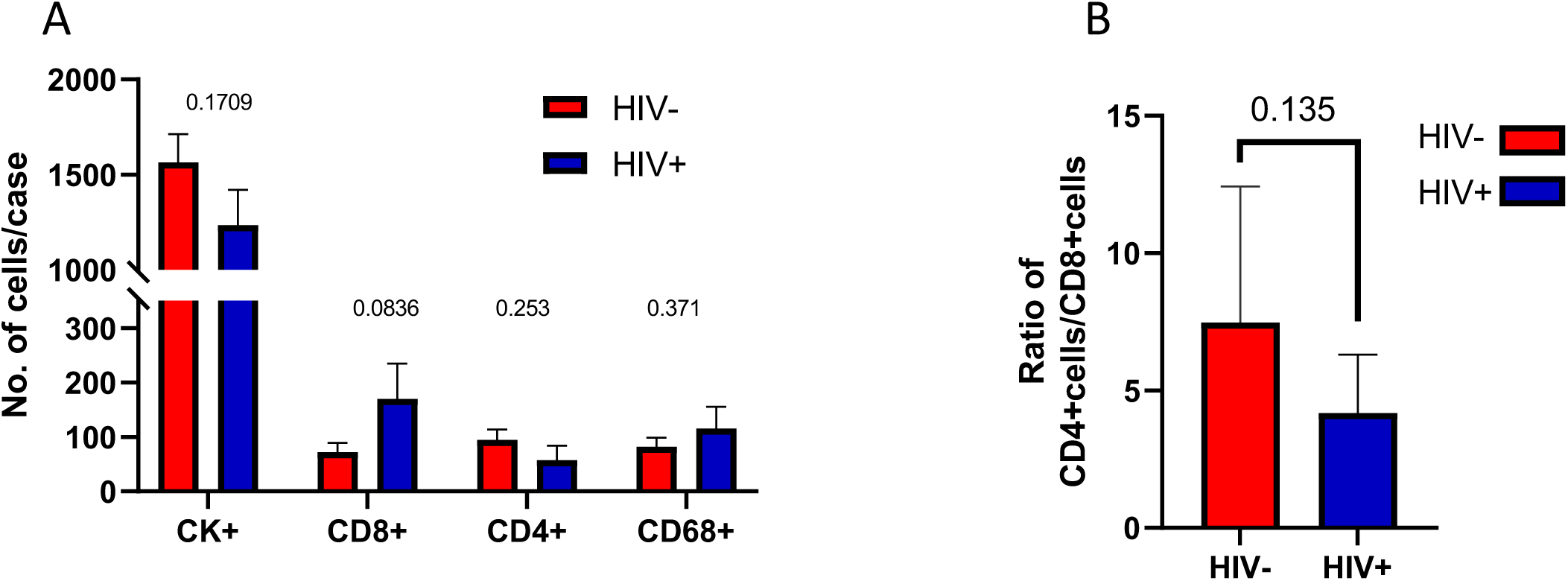
(A) Mean number of CK+ tumor cells, CD8+, CD4+ tumor-infiltrating lymphocytes (TIL) and CD68+ tumor-associated macrophages (TAM) in non-HIV controls (red bars) and in HIV+ NSCLCs (blue bars). Each bar depicts the mean±SEM (B) CD4/CD8+ TIL ratio among control uninfected NSCLC cases (red bar) and HIV-associated NSCLC cases (blue bar). Each bar depicts the mean±SEM

IMC findings were confirmed with quantitative immunofluorescence (QIF) staining and analysis which also noted no overall differences between CD4+ and CD8+ T cells or B cell infiltration into the tumor environment (data not shown). Of note, compared to non-HIV tumors, individuals with uncontrolled HIV replication (viral load >400 copies/μL) had decreased TIL for CD4 (*P* = 0.02) and CD20 (*P* = 0.02) populations. Among patients with viral suppression on antiretroviral therapy, there was a wide range of expression without a statistically significant difference compared to non-HIV cases. Among HIV+NSCLC, age, stage, AIDS status, or peripheral blood CD4+ T cell count were not associated with T cell infiltration in the TME (data not shown).

### TILs display altered functional profiles in HIV-associated NSCLC

IMC single-cell data was analyzed to identify and compare salient features of CD8+ and CD4+ TIL subpopulations in tumors from PWH and PWOH. The markers included differentiation (Tbet, CD45RO), activation (CD25, PD-1 and GZB), proliferation (Ki-67) and immune inhibitory receptors associated with T-cell exhaustion (PD-1, LAG-3 and TIM-3).

As shown in **Figure 3A** and after adjusting for multiple comparisons, CD8+ T cells from HIV-associated tumors showed significantly higher levels of CD45RO and Tbet expression; increased Ki67; increased activation with CD25 expression and granzyme B (GRZB); as well as higher levels of T-cell exhaustion markers PD-1, Lag-3, and Tim-3. A random effects model, which additionally controlled for inter-patient variation, further confirmed significantly elevated levels of Ki-67 and GRZB in CD8+ TILs from HIV+ cases, and a trend towards increased PD-1 expression (**Figure 3B**).

**Figure 3.**
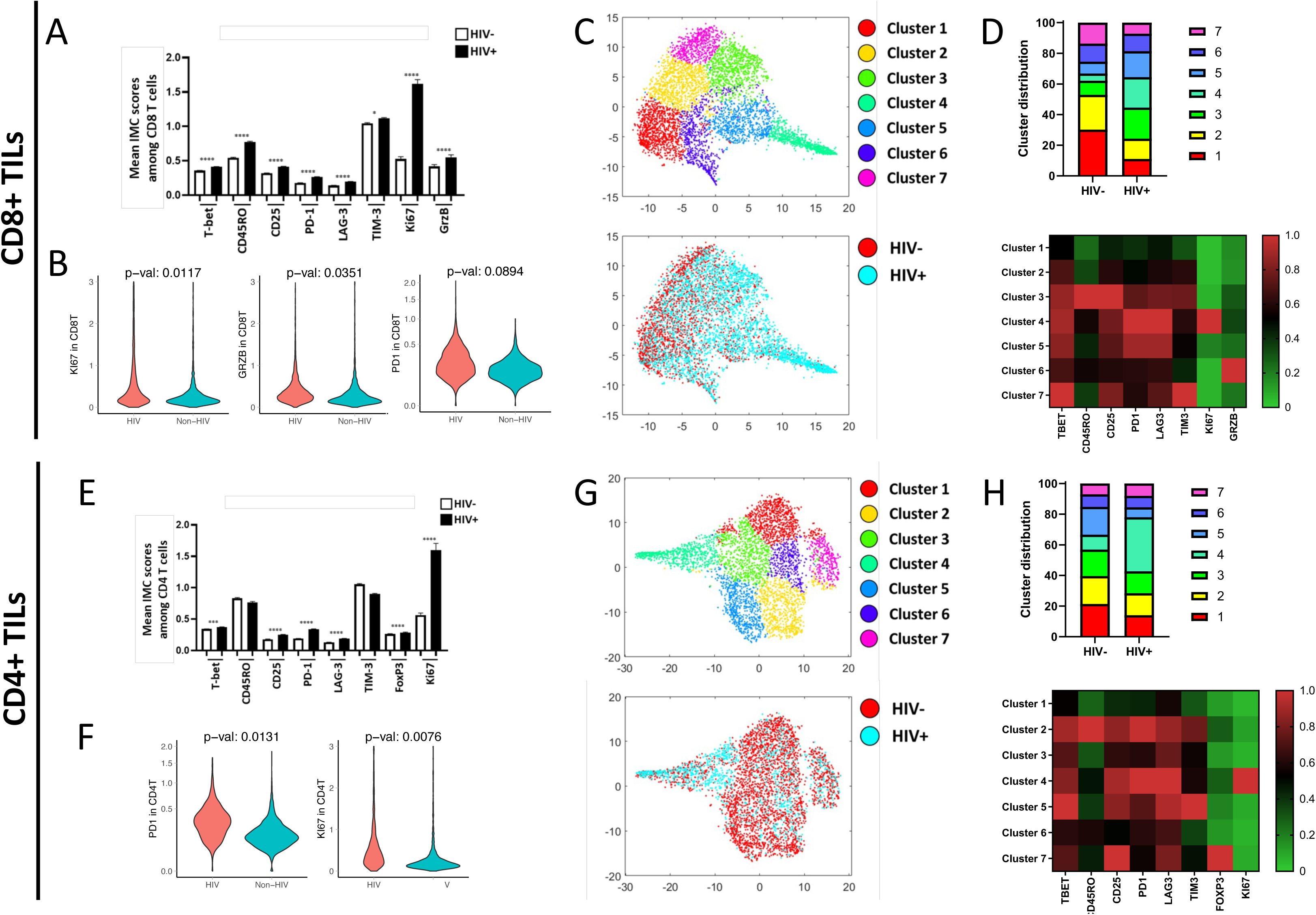
(A, D) Mean expression value of single markers within CD8+ T cells (A) and CD4+ T cells from immune mass cytometry panels. (B, E) Unsupervised clustering analysis revealing 7 unique clusters of CD8+ (B) and CD4+ (E) T cells in the top figure and distribution of cells by HIV status in bottom panel with cells from PWH (blue) and PWOH (red). (C, F) Bar graphs representing distribution of each CD8 (C) and CD4 (F) T cell cluster among PWH and PWOH. Heat map showing intensity of each marker across the different clusters.

Beyond individual marker expression which does not provide a holistic evaluation of T-cell populations, we next asked whether phenotypic multi-marker patterns of CD8+ TILs were different between HIV-associated vs. non-HIV tumors using an unsupervised clustering algorithm. Seven unique CD8+ T cell clusters were generated based on the patterns of marker expression which demonstrate a differential distribution between HIV+ and non-HIV cases (**Figure 3C**). Evaluation of the distribution of cells between HIV+ and non-HIV tumors revealed prominent differences, notably with expansion of clusters 3, 4, and 5 which together represent 57.1% of CD8+ TILs among HIV+ tumors, but only 21.7% in non-HIV tumors (p<0.001). These three clusters are unique by definition, but they each express high relative expression of PD-1 and Lag-3 comparatively, as well as lower levels of GRZB amongst all clusters. Cluster 3 is notable for very high levels of CD45RO and CD25, suggestive of an “activated effector memory cell subset”. Cluster 4 strongly resembles previously described dysfunctional “Effector burned out cells” (“Ebo” cells)[12] which have high levels of PD-1 and LAG-3, with elevated Ki-67 and low cytotoxic (GRZB) activity. Increased presence and expansion of these dysfunctional “Ebo” cells have been recently associated with limited clinical benefit from PD-1 axis blockers in patients with advanced NSCLC. Here, we show that these “Ebo” cells are even further expanded among tumors from PWH compared to PWOH (19.9% of all CD8+ T cells among HIV+ tumors, whereas this cluster comprises only 4.7% of CD8+ TILs among uninfected controls, p<0.001). Cluster 5, which also appears “highly exhausted but non-proliferating” with elevated expression of PD-1 and Lag-3 closely resembles “Ebo” cells but without elevated Ki67 expression. Of note, among non-HIV tumors, the majority of CD8+ TILs lie within Clusters 1 and 2 (52.9% of total) compared to only 24.3% among the HIV+ tumors. Clusters 1 and 2 are notable for their relative low expression of PD-1 and Lag-3, as well as higher expression of CD45RO, suggesting that these cells are less differentiated and representing a more functional central memory phenotype.

Single cell expression levels of phenotypic and functional markers studied among CD4+ TILs (**Figure 3E**) showed similar patterns to that seen among CD8+ TILs with higher levels of the activation markers CD25 and Tbet; T-cell inhibitory receptors PD-1 and Lag-3; as well as proliferation marker Ki-67 among HIV-associated tumors compared to non-HIV tumors. Notably, unlike CD8+ TILs, there was no difference in CD45RO or Tim-3 expression between the two groups. In a random effects model, PD-1 and Ki67 remained significantly elevated among CD4+ T cells from HIV+, compared with non-HIV, tumors (**Figure 3F**).

Unsupervised clustering reveals seven unique clusters which, like in the CD8+ T cell population, show differential distribution between HIV+ and non-HIV tumors (**Figure 3G, H**). Most striking is the finding that cluster 4 is most predominant among CD4+ T-cells from HIV+ tumors making up 35.2% of all CD4+ TILs in the TME, whereas among non-HIV tumors, it only makes up 9.8%. Detailed characterization of Cluster 4 reveals again very high expression of PD-1, Lag-3, as well as Ki-67, suggesting a dysfunctional profile as recently reported [19–21]. Thus, this phenotype mirrors the “Ebo” phenotype seen in CD8+ T-cells and further exhibits elevated CD25 suggesting an activated state. Cluster 7 likely represents the T regulatory cell population, based on high FoxP3 and CD25 expression, and is found in comparable proportion in both HIV+ and non-HIV tumors, at 8.0% and 7.0% respectively, despite single marker expression of FoxP3 being slightly higher among HIV+ CD4+ T-cells.

### Tumor-associated macrophages show distinct immunoregulatory features in HIV+ tumors

Tumor-associated macrophages (TAM) are recognized as increasingly important in the orchestration of anti-tumor immunity in the TME. Evaluation of CD68+ TAMs revealed differential findings between HIV-associated and non-HIV tumors. When evaluating individual markers, HIV-associated tumors have increased immunoregulatory receptors PD-L1, PD-L2, B7H3, B7H4 and VISTA (Figure 4B). In addition, TAMs in HIV-associated tumors have evidence of increased IDO1, an enzyme that converts tryptophan to kynurenine, a potent immunosuppressive molecule. And, as with T cells, TAMs in HIV-associated tumors have significantly increased Ki67 expression. These findings point to HIV-associated NSCLCs harboring TAMs that are associated with universal upregulation of markers associated with an immunoregulatory phenotype, known to render T cells less functional. Unsupervised clustering of TAMs by cumulative expression of multiple markers also demonstrates that TAMs from HIV+ vs non-HIV tumors are distinct (Figure 4C), with clusters 1, 6, and 7 making up 58.8% of TAMs from HIV-associated tumors but only 17.8% of TAMs from non-HIV tumors. Clusters 6 and 7 are notable for co-expression of multiple markers from the B7 family including PD-L1, PD-L2, B7H3, and B7H4; whereas cluster 1 is notable for high expression of VISTA. This is in stark contrast to TAMs from non-HIV tumors where clusters 2, 3, 4, and 5 make up 82.2% of TAMS (figure 4C), and these clusters are notable for lower expression of each of the immunoregulatory molecules identified.

**Figure 4.**
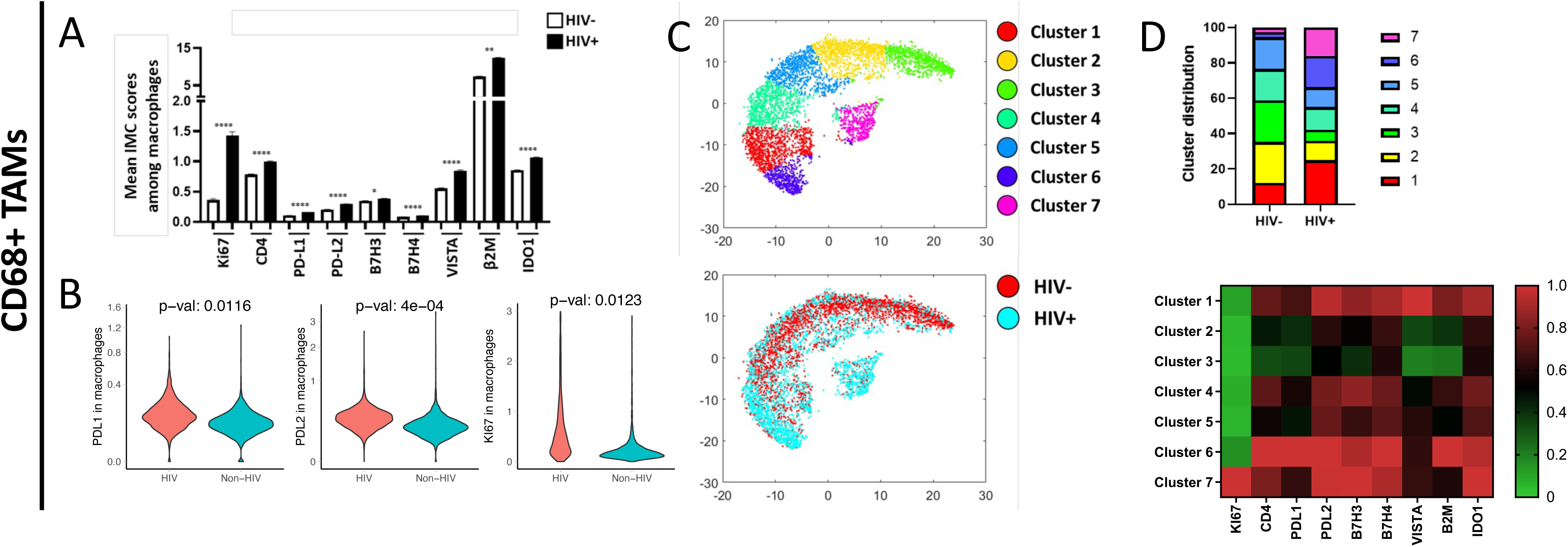
(A) Mean expression value of single markers within CD68+ tumor associated macrophages (TAMs) from immune mass cytometry panels. (B) Unsupervised clustering analysis revealing 7 unique clusters of TAMs in the top panel and distribution of cells by HIV status in bottom panel with cells from PWH (blue) and PWOH (red). (C) Bar graphs representing distribution of each TAM cluster among PWH and PWOH. Heat map showing intensity of each marker across the different clusters.

### Spectral Graph Theory confirms ability to discriminate the tumor microenvironment of HIV+ NSCLC from non-HIV NSCLC

Cell segmentation analysis algorithms are critically important for the identification of potential novel cell subsets within complex datasets such as IMC. However, in order to confirm our AI-based cell segmentation and quantitative platforms, we additionally employed an alternative approach to imaging analysis. A computational strategy based on the PageRank mathematical algorithm was used in order to establish an unsupervised and cell segmentation-independent signature associated with HIV status[22]. First, 14 individual 35 mm^2^ micro-image patches were selected per case using the highest expression of lineage-defining cell markers (CD4, CD8, and CK cells). Within each patch, tumor and neighboring immune cells were mapped with consideration of all markers. For each patch, steady-state distribution (SSD) vectors were computed, organized into a co-variance matrix, subjected to nonlinear dimensionality reduction using diffusion maps and classified using an RBF support vector machine (SVM) classifier. Finally, a leave-one-case-out cross-validation analysis was performed to show accuracy for associations with group. The integrated spatial classifier was applied to the cohort of HIV and non-HIV cases. Figure 5A demonstrates the clustering of cases where each point identifies a single patch, color-coded by HIV status. The corresponding confusion matrix was attained by a classifier applied to the low-dimensional representation obtained by diffusion maps. By training and testing the classifier as described above, HIV+ NSCLC could be distinguished from non-HIV NSCLC, providing discrimination with 84.6% accuracy (Figure 5B). The proteins most likely to differentiate HIV+ from non-HIV in rank order were PD-L2, CD25, β2M, Vimentin and PD-1 (Figure 5C). The majority of markers associated with differentiating HIV+ vs non-HIV tumors using spectral graph theory are the same markers that differentiated individual immune subsets (PDL2 expression in TAMS, PD-1 ad CD25 in T cells) using the cell segmentation approach to IMC analysis. This concordance, utilizing two different algorithms and approaches to measure complex IMC data provides confidence that expression levels of these specific markers do in fact discriminate the TME of HIV-associated vs. non-HIV NSCLC. In addition, some markers found on CK+ tumor cells including β2-microglobulin and vimentin aided in discriminating HIV vs. non-HIV associated NSCLC cells, raising future areas to explore which include differences in protein expression of tumor cells among PWH compared to PWOH.

**Figure 5.**
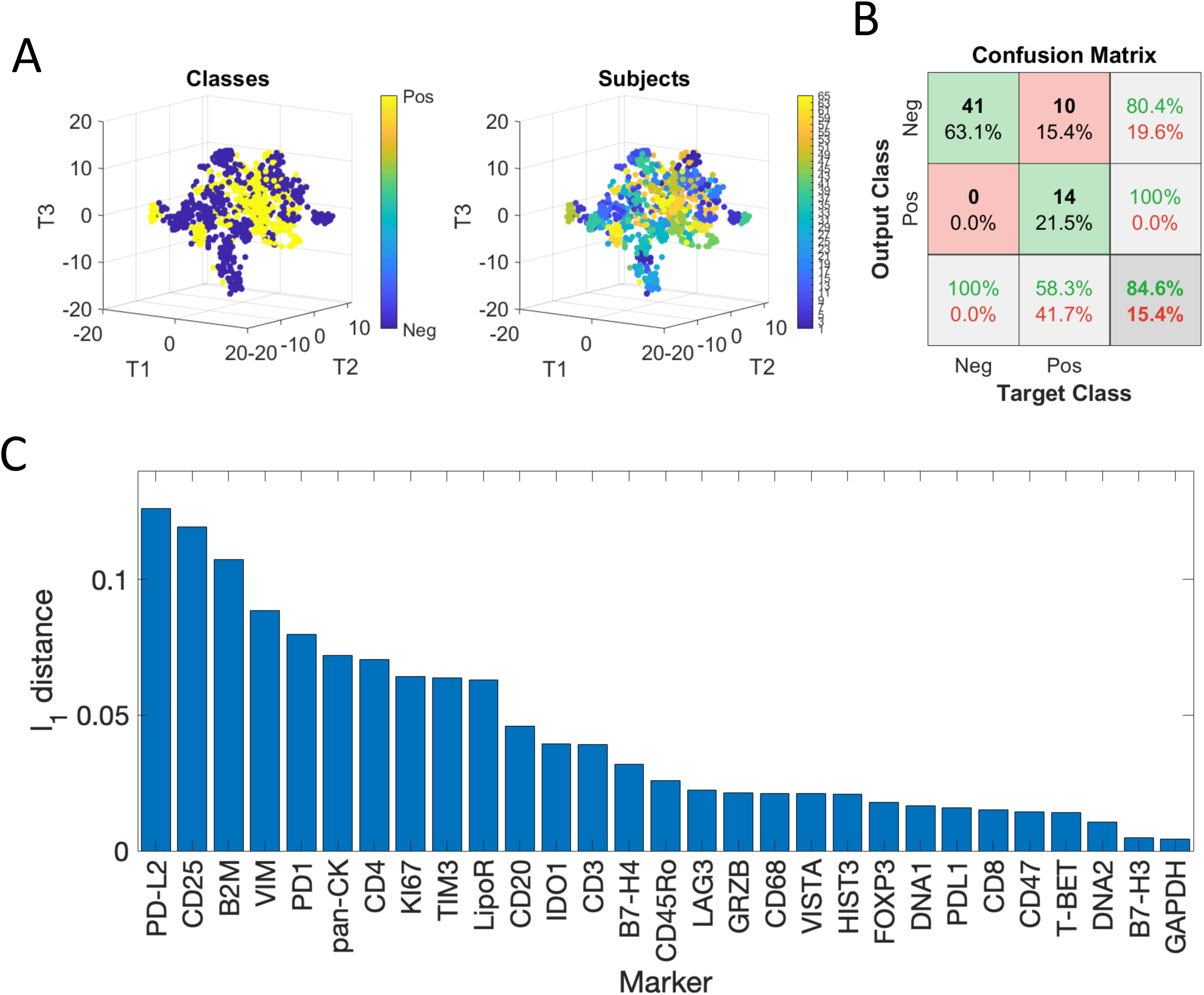
(A) Visualization of patches representation in 3D using t-SNE, colored according to (left) the HIV status and (right) the subjects. (B) The confusion matrix for HIV prediction obtained using a leave-one-subject-out cross-validation based on a Radial Basis Function (RBF) Support Vector Machine (SVM)classifier. (C)Significance of antibodies ranked according to the L1 distance between their steady-state distributions (SSD) on the graph of 14 patches associated with the highest CK expressions.

### Spatial Architecture reveals increased distances between immune cells in the HIV+ tumor microenvironment

An attractive feature of imaging mass cytometry, in additional to the ability to use a large number of phenotypic and functional markers for cell characterization, is the ability to maintain spatial architecture. The distribution of immune cells within and surrounding tumor cells is of increasing interest and has been noted to be associated with prognosis in some studies. Using our IMC data which preserves spatial architecture of the TME, we calculated distances between individual immune cell populations (CD4+ and CD8+ TILs, and CD68+ TAMs), as well as distances between those immune cell populations and tumor cells (Figure 6A). Using a method that measured the distance from every cell of one subset to each cell of a different subset, we were able to measure the average distances for each interaction tested. Of note, for every interaction measured (CD4+ to CD8+, CD4+ to tumor, CD8+ to tumor, TAM to tumor, TAM to CD4+, and TAM to CD8+), the average distances between cells was greater within HIV+ tumors than non-HIV tumors, despite similar numbers of cells within each cell population (**Figure 6B, Supplemental Table S2**). In actual distances, the cell populations that displayed the largest distance are between TAMs and all other cell types (CD4+ and CD8+ T cells, and CK+ tumor cells), as well as the distance between CD4+ and CD8+ cells. The latter is likely reflected by the lower CD4:CD8 ratio among HIV-associated tumors.

**Figure 6.**
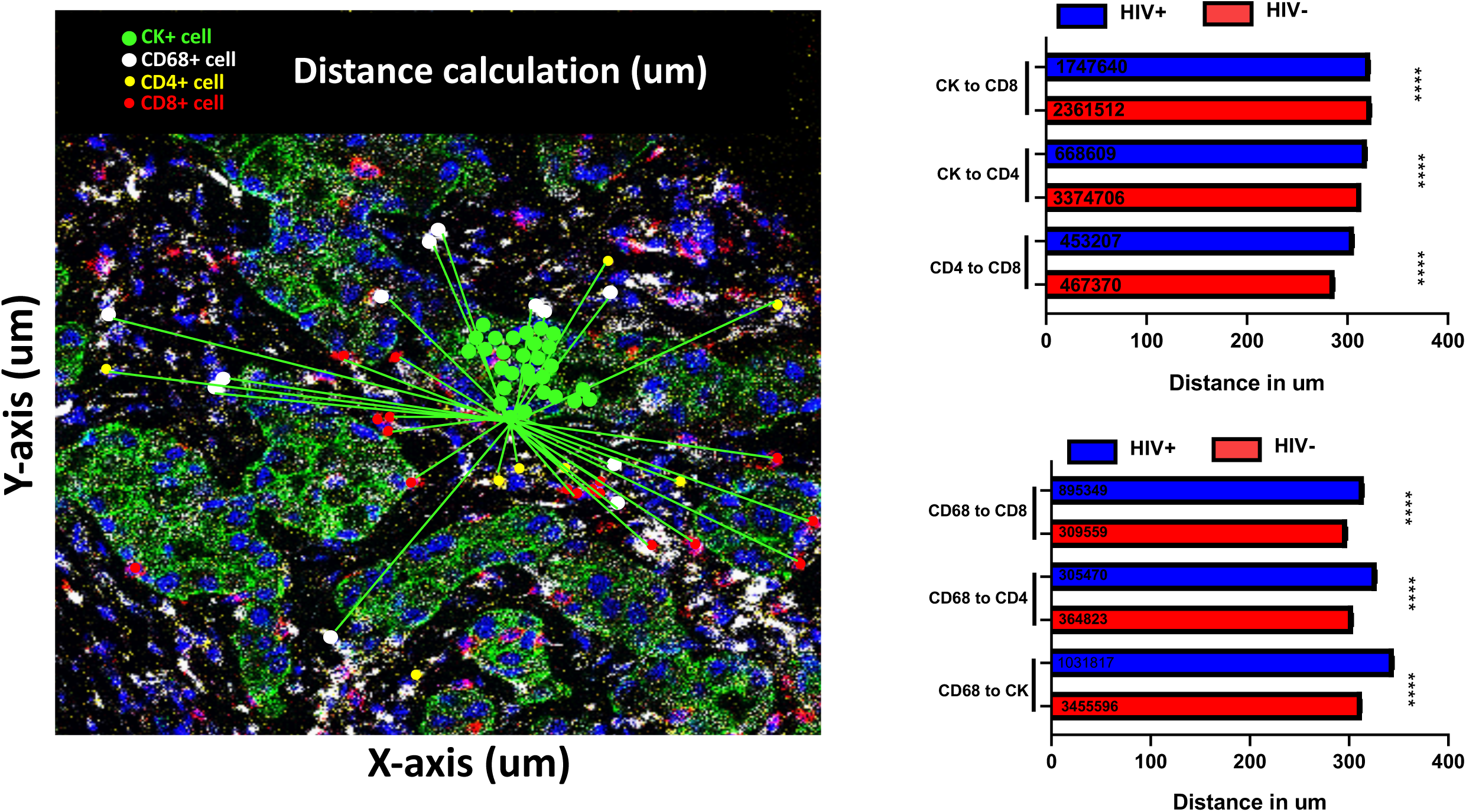
(A) Representative sample in which distances were calculated between individual cell populations (B) Average distance between cells of each phenotype with each cell of another phenotypic subset. Welch’s t-Test was performed considering different sd values. P values were corrected by Bonferroni.

### T cell mediated tumor killing is impaired in PWH compared to PWOH

Tumor cells from tumor cell line PC9 (HLA-A2+ EGFR mutant NSCLC) were either mixed with PBMCs in their native state or after activation with IFNγ and TNFα to increase antigen presentation. Tumors were then exposed to PBMCs from donors at a 2:1 or 5:1 ratio (PBMC: tumor). After three days of co-incubation, tumor killing was measured by annexin staining on EPCAM+ (tumor) cells and exhaustion was measured by Lag-3 expression on CD8+ T cells. Upon exposure to tumor cells, T cells from PWH demonstrated significantly increased exhaustion marker (Lag3) expression but decreased activation marker expression (CD25) compared to PWOH (Fig 7A). This difference was noted when co-cultured with tumor cells in their native state or after activation with IFNg and TNFa. Striking, tumor cells in the presence of PBMCs from PWH had significantly lower expression of Annexin V, a measure of cell death, compared to tumor cells cultured in the presence of PBMCs from PWOH. (**Figure 7B**). When cultured with PBMCs from PWOH, there was a dose dependent increase in tumor cell killing (i.e., more Annexin V staining with 1:5 tumor:effector cell ratio. Among PWH, increase in ratio of effector cells did not improve tumor killing, whether or not tumors were in their native state, or stimulated with cytokines to increase antigen presentation. These findings suggest that cells from PWH, despite viral suppression, exhibit increased susceptibility to T cell exhaustion when encountering a neoantigen, and further exhibit deficiencies in tumor killing. Similar findings were demonstrated with a second tumor cell line A549 that is an HLA-A2+ KRAS mutant NSCLC cell line **(Supplemental Figure 1**)

**Figure 7.**
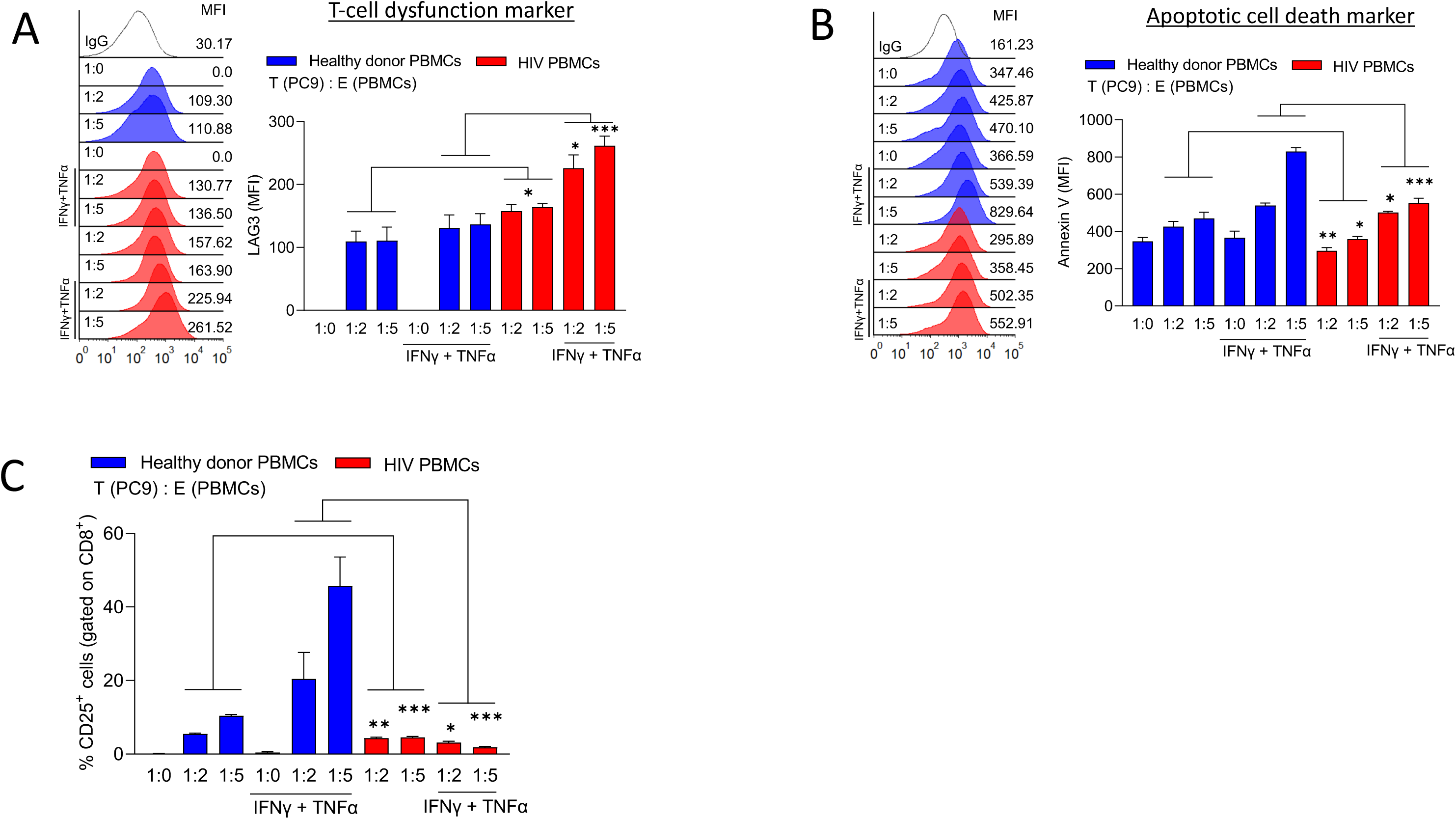
PBMCs from patients with HIV (red) and without HIV (green) were incubated with tumor cells from tumor cell PK9, an HLA-A2+ EGFR mutant NSCLC tumor cell line, that were either in their native state or stimulated for 24 hours with IFNg and TNFa. Tumor:effector T cell ration was 1:0, 1:2, or 1:5. Y-axis represents the proportion of cells that expressed either CD25 on CD8+ T cells, Lag-3 on CD8+ T cells or Annexin V on EPCAM+ tumor cells.

## Discussion

Among people living with HIV in the US, NSCLC, a non-AIDS defining cancer, is the leading cause of cancer-related death and is increased in incidence compared to the general population[2, 3]. Immune dysfunction in the setting of HIV infection is associated with chronic inflammation, both targeted to the virus as well as widespread bystander inflammation[23, 24]. In addition, disruption of lymphatic tissue architecture, including lymph nodes, thymus and bone marrow, further impact the ability to rectify years of immunologic damage even after viral suppression with antiretroviral therapy (ART)[25–27]. Global ongoing immune activation and immune dysfunction have also been associated with increased all-cause mortality, and specifically with increased cancer incidence [28–30]. However, the impact of HIV-associated immunologic changes on the tumor immune environment has not been studied and may explain the clinical finding of worse cancer-associated prognosis among PWH. This may be particularly true for those malignancies, including NSCLC, where we know that the immune response is an important indicator of prognosis. Using a well-matched cohort of NSCLC tumor tissue from PWH and PWOH, and immune mass cytometry analysis with preserved tissue architecture, we find that PWH harbor a tumor immune microenvironment (TME) that is consistent with highly exhausted, dysfunctional CD4+ and CD8+ T cells. Further, tumor associated macrophages (TAMs) exhibit a program consistent with fostering a highly immunoregulatory environment. These findings demonstrate clear differential impact of HIV infection on the immune microenvironment. Importantly, we have shown that T cells from PWH have a lower threshold to upregulate immunoregulatory receptor Lag-3, and strikingly less able to induce tumor killing of HLA-matched tumor cells, demonstrating that PWH, even after viral suppression, harbor both phenotypic and functional differences that can impact response to neoantigens. Overall, both circulating immune cells and those within the tumor tissue of patients with HIV demonstrate clear evidence of fostering a regulatory immune environment, consistent with impaired anti-tumor immunity.

The TME is an important niche where tumor cells and surrounding immune and stromal cells provide bidirectional signaling that establishes either a tumor-promoting or regulatory environment. Though the number of tumor infiltrating lymphocytes (TILs), particularly effector CD8+ T cells into the TME, has been associated with improved survival in several cancers including NSCLC[31], TIL numbers alone do not explain differences between HIV-associated and non-HIV NSCLC given the comparable levels of CD4+, CD8+, and TAM T cell infiltration seen in our study. In fact, there was a trend of increased CD8+ T cells within HIV-associated NSCLC. However, a decreased CD4:CD8 ratio within the TME suggests that decreased help from CD4+ T cells for CD8+ effector cells may be a specific regulatory factor within HIV-associated tumors, as it is in the circulation. CD4:CD8 ratio is inverted early after HIV infection and its persistence after ART has been associated with increased cancer incidence and all-cause mortality, emphasizing this specific feature of HIV-associated immunologic perturbation as impactful in the anti-tumor responses[32, 33].

In addition, we found that CD8+ T cells within the TME from PWH demonstrate a dysfunctional phenotype, with increased expression of PD-1 and other inhibitory receptors (e.g. Lag-3, Tim-3) on T cells rendering them less able to perform cytotoxic killing. Immune checkpoint blockers (ICBs), such as those targeting the PD-1:PD-L1 pathway have been used with significant improvements in outcome for some patients[34]. However, the heterogeneity of expression of inhibitory receptors among the TME among patients have demonstrated that those patients who harbor T cells particularly “dysfunctional” or “exhausted cells” to be less likely to response to ICBs. Recently, a cell-subtype referred to as “Effector burned out” (Ebo) cells were described, reflecting an apoptosis-resistant, proliferative, but dysfunctional T cell population with evidence of multiple inhibitory receptors including PD-1, Lag-3, and Tim-3[35]. The presence of Ebo cells has been associated with poor response to ICB therapies. In this study, we find that HIV+ tumors harbor striking increases in “Ebo cells” among both CD4+ and CD8+ subsets, compared to non-HIV tumors. Accumulation of effector-like, apoptosis-resistant CD8+ T cells have been well-described in the peripheral blood of HIV infection, secondary to chronic hyper-inflammatory state. We surmise that this global phenomenon seen among PWH is thus further accentuated in the TME, allowing tumors to grow with decreased immune pressure.

Parallel to changes in the adaptive immune responses (i.e. T cells), TAMs from HIV-associated tumors demonstrate some of the most striking differences compared to non-HIV tumors. CD68+ TAMs from PWH display multiple ligands from the B7 family, including PD-L1 (B7-H1), PD-L2 (B7-H2), B7-H3, and B7-H4, which bind to receptors on T cells regulating their cytolytic and cytotoxic responses. Expression of PD-L2 is particularly elevated across TAMs from HIV-associated tumors and is, in fact, the primary marker identified using spectral graph theory that distinguishes HIV-associated tumors compared to non-HIV tumors. This finding raises the possibility of exploring PD-L2 as a potential target in HIV+ tumors. In addition, TAMs from HIV+ tumors also overexpress VISTA, another immune checkpoint as well as IDO1, an enzyme associated with metabolic changes associated with immunoregulation. Though PD-L1, a current target of several immune checkpoint inhibitors used clinically, is also increased among HIV-associated TAMs, the highly increased expression across other members of the B7 family, as well as VISTA and IDO1, suggests that anti-PDL1 strategies may be less effective among PWH due to multiple, overlapping pathways by which regulatory signaling to T cells is augmented, further contributing to poor anti-tumor immunity.

The role of immune mass cytometry as a novel methodology with simultaneous measurement of multiple markers as well allows for dynamic characterization of the TME. Using both cell segmentation and segmentation-independent approaches, we have concordance in the findings that HIV-associated NSCLC has evidence of a more immunoregulatory landscape. Artificial Intelligence (AI)-based approaches were also able to differentiate between HIV-associated and uninfected tumors with >80% accuracy in this small dataset, with PDL2 (on TAMS) and CD25 (on T cells) respectively being the most discriminating features of an HIV-associated TME. These analyses suggest that targeting immune cells in the TME, an approach that is increasingly being embraced in drug development, may require specific strategies among PWH where the differences in immune cells are distinct. Further refinement of markers and larger datasets may result in even more robust differentiation between these two groups.

The finding that T cells from PWH, even after durable viral suppression on ART, have a higher likelihood of upregulating immune regulatory receptors, and decreased capacity for tumor killing, provides further evidence that circulating immune cells among PWH have decreased cytotoxicity, likely due to decreased thresholds to upregulate markers of exhaustion. This functional defect is likely to affect the ability of PWH to respond to neoantigens in vivo, including in the tumor environment of malignancies like NSCLC, which we know are dependent on an effective immune response for control of tumor growth.

Using a cohort of PWH and PWOH with NSCLC from single institution, matched for stage, histology, age, tobacco use, and year of diagnosis, we performed a comprehensive evaluation of TME among HIV-NSCLC using multiple imaging modalities and bioinformatic analytic platforms, this study for the first time systematically studied the diversity of the TME among NSCLC among PWH and uninfected well-matched controls. Our findings suggest that the TME of HIV-associated NSCLC is fundamentally different from non-HIV NSCLC, and that circulating T cells from PWH, even after effective HIV control, demonstrate decreased ability to kill tumor cells. These findings are critically important for a patient population at higher risk for NSCLC who present at a younger age and suffer worse prognosis. Importantly, it identifies unique targets for this patient population that requires further evaluation with larger cohort studies. These differences also highlight the need for PWH to be included in clinical trials for ICI therapies in order to evaluate responses in this unique patient population. Similarity of TME features among those with HIV-associated NSCLC and those who respond poorly to ICI in the general population further suggests that HIV-NSCLC may further illuminate features of NSCLC that reflect non-response to current immune-based approaches.

## Data Availability

All data produced in the present study are available upon reasonable request to the authors.

## REFERENCES

1. Robbins, H.A., et al., Excess cancers among HIV-infected people in the United States. J Natl Cancer Inst, 2015. 107(4).

2. Engels, E.A., et al., Cancer-Attributable Mortality Among People With Treated Human Immunodeficiency Virus Infection in North America. Clin Infect Dis, 2017. 65(4): p. 636–643.

3. Winstone, T.A., et al., Epidemic of lung cancer in patients with HIV infection. Chest, 2013. 143(2): p. 305–314.

4. Hessol, N.A., et al., Lung cancer incidence and survival among HIV-infected and uninfected women and men. AIDS, 2015. 29(10): p. 1183–93.

5. Hleyhel, M., et al., Risk of non-AIDS-defining cancers among HIV-1-infected individuals in France between 1997 and 2009: results from a French cohort. AIDS, 2014. 28(14): p. 2109–18.

6. Marcus, J.L., et al., Immunodeficiency, AIDS-related pneumonia, and risk of lung cancer among HIV-infected individuals. AIDS, 2017. 31(7): p. 989–993.

7. Hysell, K., et al., Decreased Overall Survival in HIV-associated Non-small-cell Lung Cancer. Clin Lung Cancer, 2021. 22(4): p. e498–e505.

8. Sokoya, T., et al., HIV as a Cause of Immune Activation and Immunosenescence. Mediators Inflamm, 2017. 2017: p. 6825493.

9. Schalper, K.A., et al., Objective measurement and clinical significance of TILs in non-small cell lung cancer. J Natl Cancer Inst, 2015. 107(3).

10. Datar, I., et al., Expression Analysis and Significance of PD-1, LAG-3, and TIM-3 in Human Non-Small Cell Lung Cancer Using Spatially Resolved and Multiparametric Single-Cell Analysis. Clin Cancer Res, 2019. 25(15): p. 4663–4673.

11. Gettinger, S.N., et al., A dormant TIL phenotype defines non-small cell lung carcinomas sensitive to immune checkpoint blockers. Nat Commun, 2018. 9(1): p. 3196.

12. Sanmamed, M.F., et al., A burned-out CD8+ T-cell subset expands in the tumor microenvironment and curbs cancer immunotherapy. Cancer Discov, 2021.

13. Caushi, J.X., et al., Transcriptional programs of neoantigen-specific TIL in anti-PD-1-treated lung cancers. Nature, 2021. 596(7870): p. 126–132.

14. Lopez de Rodas, M., et al., Role of tumor infiltrating lymphocytes and spatial immune heterogeneity in sensitivity to PD-1 axis blockers in non-small cell lung cancer. J Immunother Cancer, 2022. 10(6).

15. Mandarano, M., et al., Assessment of TILs, IDO-1, and PD-L1 in resected non-small cell lung cancer: an immunohistochemical study with clinicopathological and prognostic implications. Virchows Arch, 2019. 474(2): p. 159–168.

16. Mlecnik, B., et al., Histopathologic-based prognostic factors of colorectal cancers are associated with the state of the local immune reaction. J Clin Oncol, 2011. 29(6): p. 610–8.

17. Bonaventura, P., et al., Cold Tumors: A Therapeutic Challenge for Immunotherapy. Front Immunol, 2019. 10: p. 168.

18. Levine, J.H., et al., Data-Driven Phenotypic Dissection of AML Reveals Progenitor-like Cells that Correlate with Prognosis. Cell, 2015. 162(1): p. 184–97.

19. Fu, J., et al., CD4(+) T cell exhaustion leads to adoptive transfer therapy failure which can be prevented by immune checkpoint blockade. Am J Cancer Res, 2020. 10(12): p. 4234–4250.

20. Nagasaki, J., et al., The critical role of CD4+ T cells in PD-1 blockade against MHC-II-expressing tumors such as classic Hodgkin lymphoma. Blood Adv, 2020. 4(17): p. 4069–4082.

21. Rad Pour, S., et al., Exhaustion of CD4+ T-cells mediated by the Kynurenine Pathway in Melanoma. Sci Rep, 2019. 9(1): p. 12150.

22. Lin, Y.E., et al., Graph of graphs analysis for multiplexed data with application to imaging mass cytometry. PLoS Comput Biol, 2021. 17(3): p. e1008741.

23. Emu, B., et al., Composition and function of T cell subpopulations are slow to change despite effective antiretroviral treatment of HIV disease. PLoS One, 2014. 9(1): p. e85613.

24. Boyd, M.A., et al., Navigating the complexity of chronic HIV-1 associated immune dysregulation. Curr Opin Immunol, 2022. 76: p. 102186.

25. Estes, J.D., Pathobiology of HIV/SIV-associated changes in secondary lymphoid tissues. Immunol Rev, 2013. 254(1): p. 65–77.

26. Zeng, M., A.T. Haase, and T.W. Schacker, Lymphoid tissue structure and HIV-1 infection: life or death for T cells. Trends Immunol, 2012. 33(6): p. 306–14.

27. Zeng, M., et al., Lymphoid tissue damage in HIV-1 infection depletes naive T cells and limits T cell reconstitution after antiretroviral therapy. PLoS Pathog, 2012. 8(1): p. e1002437.

28. Chaudhary, O., et al., Patients with HIV-associated cancers have evidence of increased T cell dysfunction and exhaustion prior to cancer diagnosis. J Immunother Cancer, 2022. 10(4).

29. Silverberg, M.J., et al., Risk of cancers during interrupted antiretroviral therapy in the SMART study. AIDS, 2007. 21(14): p. 1957–63.

30. Sigel K, E.B., Tate JP, Dubrow R, Justice AC, Low CD4/CD8 Ratio as a Predictor of Cancer Risk in HIV-Infected Persons. CROI 2016. Boston.

31. Reynders, K. and D. De Ruysscher, Tumor infiltrating lymphocytes in lung cancer: a new prognostic parameter. J Thorac Dis, 2016. 8(8): p. E833–5.

32. Sigel, K., et al., Immunological and infectious risk factors for lung cancer in US veterans with HIV: a longitudinal cohort study. Lancet HIV, 2017. 4(2): p. e67–e73.

33. Serrano-Villar, S., et al., Increased risk of serious non-AIDS-related events in HIV-infected subjects on antiretroviral therapy associated with a low CD4/CD8 ratio. PLoS One, 2014. 9(1): p. e85798.

34. Alexander, M., S.Y. Kim, and H. Cheng, Update 2020: Management of Non-Small Cell Lung Cancer. Lung, 2020. 198(6): p. 897–907.

35. Sanmamed, M.F., et al., A Burned-Out CD8(+) T-cell Subset Expands in the Tumor Microenvironment and Curbs Cancer Immunotherapy. Cancer Discov, 2021. 11(7): p. 1700–1715.

