## Supplemental Tables and Figures for "Spatial Analysis Reveals Impaired Immune Cell Function within the Tumor Microenvironment of HIV-associated Non-small Cell Lung Cancer"

### Supplemental Table 1: Patients and Demographics

| Patient Characteristics | HIV-NSCLC N=18 | | Uninfected N=19 | | |
|---|---|---|---|---|---|
| | N | % | N | % | P-value |
| Median age | 54.1 | | 57.2 | | 0.22 |
| Gender | | | | | 0.06 |
| Male | 14 | 78% | 9 | 47% | |
| Female | 4 | 22% | 10 | 53% | |
| Smoking Status | | | | | 0.49 |
| Non-smoker | 0 | 0% | 2 | 11% | |
| Smoking History | 18 | 100% | 17 | 89% | |
| Year of Diagnosis | | | | | 0.7 |
| 1999-2004 | 4 | 22% | 6 | 32% | |
| 2005-2010 | 8 | 44% | 6 | 32% | |
| 2011-2016 | 6 | 33% | 7 | 37% | |
| Histology | | | | | 0.77 |
| Adenocarcinoma | 8 | 44% | 10 | 53% | |
| Squamous Cell | 8 | 44% | 8 | 42% | |
| NSCLC (unspecified) | 2 | 11% | 1 | 5% | |
| Stage at diagnosis | | | | | 0.96 |
| Stage I | 6 | 33% | 6 | 32% | |
| Stage II | 1 | 6% | 2 | 11% | |
| Stage III | 4 | 22% | 4 | 21% | |
| Stage IV | 7 | 39% | 7 | 37% | |
| Years of HIV Diagnosis | | | | | |
| Median | 17 | - | | | |
| Range | 2-36 | - | | | |
| AIDS Diagnosis | 8 | 44% | | | |
| HIV VL (copies/mL) | | | | | |
| <400 | 8 | 44% | | | |
| >400 | 10 | 56% | | | |
| CD4 count (cells/µL) | | | | | |
| Median | 440 | - | | | |
| Range | 26-872 | - | | | |

### Supplemental Table 2: Number of T cells within TME

| | P value | Mean (HIV) | Mean (non-HIV) | Difference | SE of difference | t ratio | df | q value |
|---|---|---|---|---|---|---|---|---|
| CK+ | 0.170988 | 1236 | 1564 | 328.8 | 237.3 | 1.386 | 60 | 0.703621 |
| CD8+ | 0.083642 | 170.2 | 72 | -98.16 | 55.54 | 1.767 | 47 | 0.697019 |
| CD4+ | 0.253303 | 57.74 | 94.84 | 37.11 | 32.1 | 1.156 | 49 | 0.703621 |
| CD68+ | 0.371457 | 115.8 | 82.43 | -33.32 | 36.97 | 0.9014 | 53 | 0.773868 |

#### Supplementary Figure S1: Tumor Killing Assay with Circulating Cells from PWH and PWOH (A549)

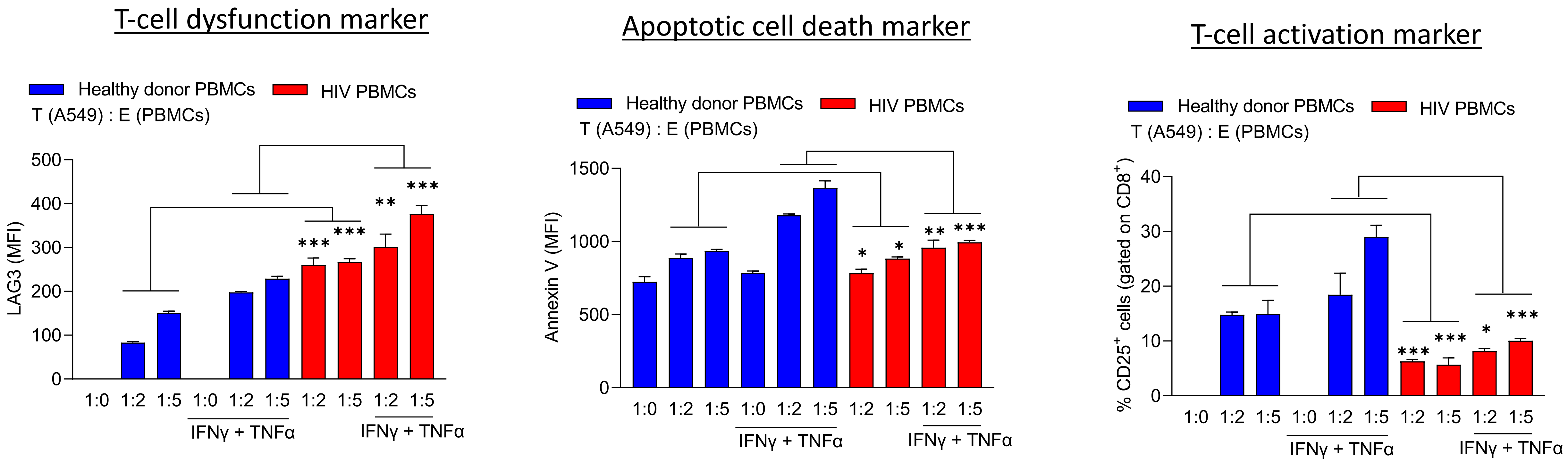
